# Artificial Intelligence methods for Improved Detection of undiagnosed Heart Failure with Preserved Ejection Fraction (HFpEF)

**DOI:** 10.1101/2023.09.12.23295413

**Authors:** Jack Wu, Dhruva Biswas, Matthew Ryan, Brett Bernstein, Maleeha Rizvi, Natalie Fairhurst, George Kaye, Ranu Baral, Tom Searle, Narbeh Melikian, Daniel Sado, Thomas F Lüscher, Richard Grocott-Mason, Gerald Carr-White, James Teo, Richard Dobson, Daniel I Bromage, Theresa A McDonagh, Ajay M Shah, Kevin O’Gallagher

## Abstract

**Background and aim:** Heart Failure with preserved Ejection Fraction (HFpEF) remains under-diagnosed in clinical practice despite accounting for nearly half of all Heart Hailure (HF) cases. Accurate and timely diagnosis of HFpEF is crucial for proper patient management and treatment. In this study, we explored the potential of natural language processing (NLP) to improve the detection and diagnosis of HFpEF according to the European Society of Cardiology (ESC) diagnostic criteria.

**Methods:** In a retrospective cohort study, we used an NLP pipeline applied to the Electronic Health Record (EHR) to identify patients with a clinical diagnosis of HF between 2010-2022. We collected demographic, clinical, echocardiographic and outcome data from the EHR. Patients were categorised according to the left ventricular ejection fraction (LVEF). Those with LVEF ≥ 50% were further categorised based on whether they had a clinician-assigned diagnosis of HFpEF and if not, whether they met the ESC diagnostic criteria. Results were validated in a second, independent centre.

**Results:** We identified 8606 patients with HF. Of 3727 consecutive patients with HF and LVEF ≥ 50% on echocardiogram, only 8.3% had a clinician-assigned diagnosis of HFpEF, while 75.4% met ESC criteria but did not have a formal diagnosis of HFpEF. Patients with confirmed HFpEF were hospitalised more frequently; however the ESC criteria group had a higher 5-year mortality, despite being less co-morbid and experiencing fewer acute cardiovascular events.

**Conclusions:** This study demonstrates that patients with undiagnosed HFpEF are an at-risk group with high mortality. It is possible to use NLP methods to identify likely HFpEF patients from EHR data who would likely then benefit from expert clinical review and complement the use of diagnostic algorithms.

**Graphical Abstract:** Of 3727 consecutive patients with a clinical diagnosis of HF and left ventricular ejection fraction (LVEF) >50% on echocardiogram, only 8.3% had a clinician-assigned diagnosis of HFpEF, while 75.4% met ESC criteria but did not have a formal diagnosis of HFpEF. The two groups had similar rates of hospitalisation however the ESC criteria group had a higher 5-year mortality.

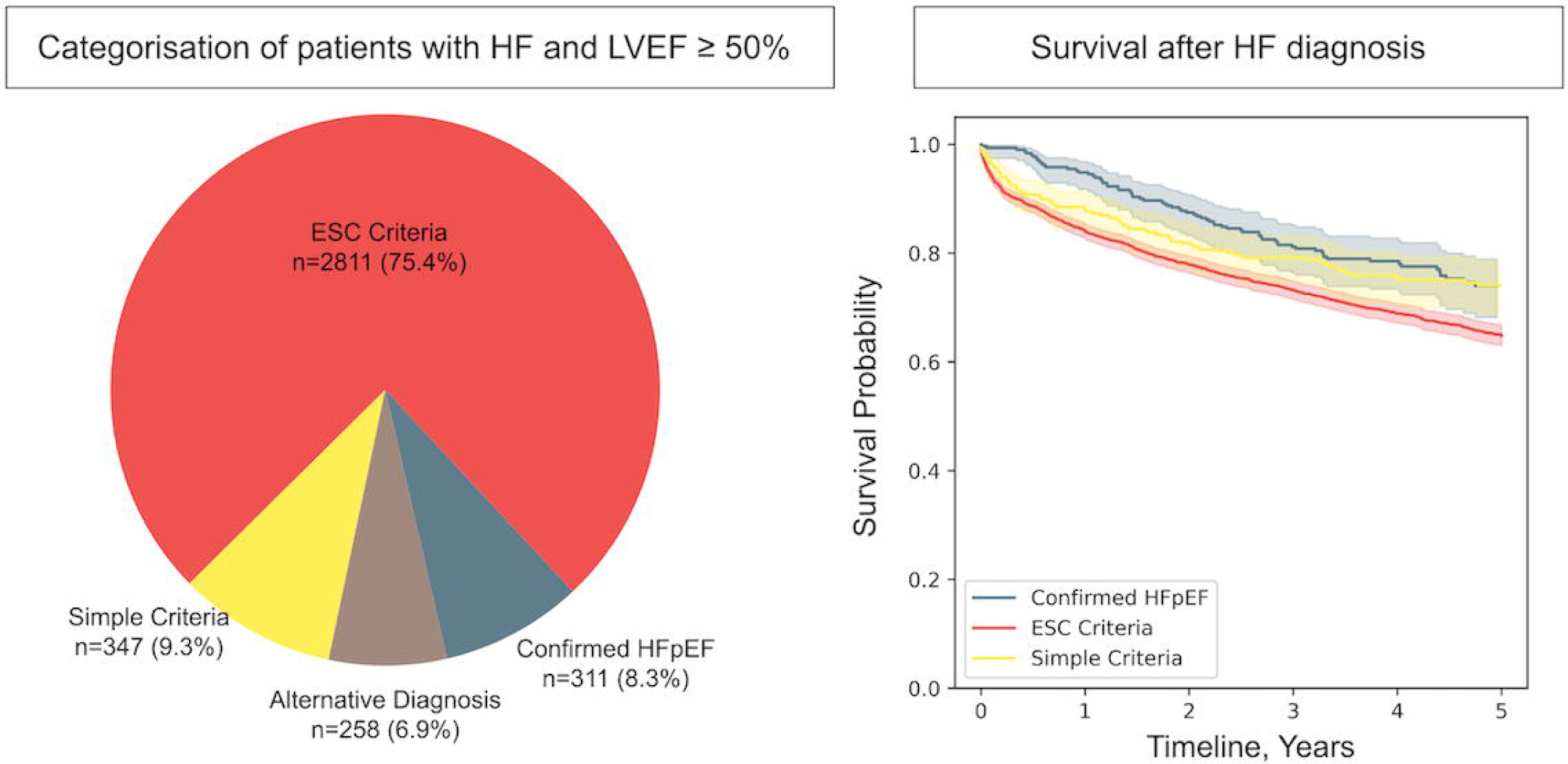

## Introduction

Heart Failure with Preserved Ejection Fraction (HFpEF) is associated with significant morbidity, healthcare utilisation, and mortality. Approximately half of all cases of Heart Failure (HF) occur in patients with preserved left ventricular ejection fraction (LVEF ≥ 50%). ^1^ In the general population over the age of 65, HFpEF is the predominant form of heart failure, and its proportion of total heart failure cases increases with age. ^2^ The HFpEF population, when compared to the populations with heart failure with reduced or mildly reduced ejection fraction (HFrEF, HFmrEF), is older, and includes a higher proportion of female patients and of those with atrial fibrillation (AF), chronic kidney disease, and non-cardiovascular co-morbidities.^3,4^

It is recognised that HFpEF can be a challenging to diagnose. Diagnostic scores such as H2FPEF and HFA-PEFF can help clinicians identify patients likely to have a diagnosis of HFpEF. ^5,6^ To reach a formal diagnosis of HFpEF by the European Society of Cardiology criteria, patients should have: 1) symptoms and signs of heart failure, 2) LVEF ≥ 50%, and 3) objective imaging or biochemical evidence of diastolic dysfunction/raised filling pressures, including left ventricular hypertrophy, left atrial dilatation, raised E/e’, and/or raised natriuretic peptides. ^4^ This is contrasted with the simplified inclusion criteria that were seen in early HFpEF therapeutic trials (although most recent clinical trials have required evidence of diastolic dysfunction/raised filling pressures for eligibility). ^7,8^ Given that HFpEF prediction scores and diagnostic criteria are relatively recent developments, it is probable that there are many patients with a prior diagnosis of HF where the formal diagnosis of HFpEF has not been made.

We hypothesised that the proportion of patients who have been formally diagnosed with HFpEF would be significantly smaller than the number of HF patients meeting ESC diagnostic criteria for HFpEF. We therefore aimed to use artificial intelligence methods – specifically natural language processing (NLP) - to identify all patients with a clinician-assigned diagnosis of HFpEF at a large London hospital and compare this cohort to a population of HF patients at the same centre who meet the ESC criteria but who remain without a formal diagnosis of HFpEF.

## Methods

### Ethics

This project operated under London South-East Research Ethics Committee approval (18/LO/2048) granted to the King’s Electronic Records Research Interface (KERRI) and London Dulwich Research Ethics Committee approval (19/LO/1957), which did not require written informed patient consent. This study complies with the Declaration of Helsinki.

### Study Design and inclusion criteria

We established a single centre retrospective anonymised database of adult patients with a clinical diagnosis of HF documented within the Electronic Health Record (EHR) at King’s College Hospital NHS Foundation Trust (KCHNFT) between 2010-2022. KCHNFT is a large, multi-site teaching hospital in London, UK providing secondary and tertiary level cardiology services and a dedicated heart failure service open to referral by any physician. Patients were included if they had two or more mentions of a “heart failure” diagnosis in the clinical text as determined by NLP.^9–12^ Patients were included regardless of inpatient or outpatient status. Both structured and unstructured portions of the echocardiogram report were used to extract LVEF data of patients and other relevant echocardiographic parameters. Patients with a clinical diagnosis of HF and LVEF ≥50% were categorised into one of the following 4 groups:

1. Confirmed HFpEF: Clinician-assigned diagnosis of HFpEF
2. Undiagnosed HFpEF (ESC Criteria): Patients with HF, LVEF≥50%, and imaging/biochemical evidence of diastolic dysfunction meeting the ESC diagnostic criteria^4^ but who have not received a HFpEF diagnosis.
3. Simple Criteria: Patients with a clinical diagnosis of HF and LVEF≥50% on echocardiography, but not meeting ESC diagnostic criteria and who have not received a HFpEF diagnosis.
4. Alternative diagnosis: This group included patients with severe valvular heart disease, hypertrophic cardiomyopathy, restrictive cardiomyopathy, constrictive pericarditis, and cardiac amyloidosis.

Patients were excluded if at any point they had an echocardiogram with LVEF <50%. Supplementary Table 2 delineates the SNOMED terms employed to identify patients for groups 1 and 4. The sequence of identification amongst the four groups proceeded as follows: Initially, Group 4 (Alternative diagnosis) was identified. Subsequently, remaining patients possessing the HFpEF term were allocated to Group 1 (Confirmed HFpEF). Utilizing the ESC criteria, patients not placed in groups 1 and 4 were assigned to either Group 2 (ESC Criteria), if they satisfied any of the criteria, or alternatively to Group 3 (Simple Criteria) if they did not meet any criteria.

A validation cohort database was constructing containing patient data from the Royal Brompton Hospital, London (Guy’s and St Thomas’ NHS Foundation Trust) using the same criteria as the KCH dataset.

### Data processing

Demographic (age, sex, self-ascribed ethnicity) and clinical data were retrieved from structured and unstructured components of the electronic health record using a variety of wellDvalidated natural language processing (NLP) informatics tools belonging to the open-source CogStack ecosystem, ^13^ MedCAT (Medical Concept Annotation Tool) ^10^ and MedCATTrainer ^14^ deployed at King’s College Hospital NHS Foundation Trust.

The CogStack NLP pipeline captures negation, synonyms, and acronyms for medical SNOMEDDCT (Systematized Medical Nomenclature for Medicine–Clinical Terminology) concepts as well as surrounding linguistic context using deep learning and long shortDterm memory networks. MedCAT produced unsupervised annotations for all SNOMEDDCT concepts under parent terms Clinical Finding, Disorder, Organism, and Event with disambiguation, preDtrained on MIMICDIII (Medical Information Mart for Intensive Care-III). ^15^ Further supervised training improved detection of annotations and metaDannotations such as experiencer (is the concept annotated experienced by the patient or other), negation (is the concept annotated negated or not) and temporality (is the concept annotated in the past or present) with MedCATTrainer. MetaDannotations for hypothetical and experiencer were merged into irrelevant meaning that any concept annotated as either hypothetical or where the experiencer was not the patient was annotated as irrelevant. Performance of the MedCAT NLP pipeline for disorders mentioned in the text has previously been evaluated on 5617 annotations for 265 documents by a domain expert (JTHT) and F1, precision and recall recorded. ^16^ For the current study, a random sample of 100 patients in the ‘Confirmed HFpEF’ group were selected for manual validation .

Using MedCAT, SNOMED-CT concepts are extracted from clinical notes of HF patients. The concepts include cardiovascular comorbidities (myocardial infarction, ischemic heart disease, cerebrovascular accident, transient ischemic attack, peripheral vascular disease, atrial fibrillation and pulmonary hypertension), non-cardiovascular comorbidities (type I/II diabetes, hypertension, kidney disease and depressive disorder), medications (calcium channel blockers, Angiotensin converting enzyme inhibitors (ACEi), angiotensin receptor blockers (ARB), beta blockers, loop diuretics, statin, metformin and also 5 drug groups: (1) Antiplatelet agents including aspirin, clopidogrel, prasugrel and ticagrelor; (2) Anticoagulants including warfarin, dabigatran, apixaban and edoxaban; (3) Mineralocorticoid receptor antagonists (MRA) including spironolactone and eplerenone; (4) Angiotensin receptor neprilysin inhibitors (ARNI) including sacubitril/valsartan; (5) Sodium-Glucose co-transporter-2 inhibitors (SGLT2i) including dapagliflozin and empagliflozin) and symptoms (dyspnoea, dyspnoea at rest, chest pain, dizziness, presyncope and syncope). Lab results are also retrieved from the CogStack pipeline; they include BMI, haemoglobin, creatinine, cholesterol, BNP and NT-proBNP. The observations are time-series data and the one closest to the first HF mention time is used as a baseline.

### Identification of ESC Criteria for evidence of diastolic dysfunction and H2FPEF score

From the EHR, we identified the ESC Criteria for evidence of cardiac structural, functional and serological abnormalities consistent with the presence of left ventricular diastolic dysfunction/raised left ventricular filling pressures.^4^ Where a criterion is indexed to body surface area (i.e. LV mass index, LV volume index) but where no indexed value could be found in the EHR, we collected data for the raw value and applied these values (i.e. LV mass in grams, LA volume in ml). The H2FPEF score was calculated for each individual patient using available data. ^5^ For criteria where alternative thresholds apply in the setting of atrial fibrillation (e.g. LA volume index, natriuretic peptides) these were applied where relevant.

### Outcome data

Mortality data were obtained from death notification letters in the EPR system and the Patient Information Management System (PIMS) of the hospital (updated from the NHS central database). Hospitalisations were estimated by the number of discharge notifications in the EHR in the study timeframe i.e. 2010-2022. Diagnoses of myocardial infarction and stroke were recorded as outcome data if they occurred after the first mention of HF in the EHR.

### Statistical analysis

All statistical tests were performed using python version 3.9.7. Mann-Whitney U test (scipy.stats package version 1.7.1) with multiple testing correction (statsmodels.stats.multitest package version 0.12.2) was used to compare the distributions of H2FPEF score and hospitalisations between the HFpEF subgroups. Kaplan Meier curves were plotted using the lifelines package version 0.27.3 and the logrank-test function in the package was used to compare the survival distributions of the HFpEF subgroups. Baseline was taken as the time of first HF diagnosis mention in the EHR. Cox proportional hazards regression with the CoxPHFitter function in the same lifelines package was used to assess relationship between patient outcome (death) and various covariates. Covariates included in the model were selected based on their clinical relevance. The number of comorbidities was included as an indicator of overall patient health status. Age at diagnosis was incorporated to control for the effects of age on survival. The number of hospitalizations and cardiology consultations were also included as measures of disease severity and healthcare utilization, respectively. Specifically, the number of cardiology consultations was quantified based on the count of cardiology letters for each patient. Lastly, sex was included as a binary covariate to account for potential differences in outcomes between males and females. The results of the Cox regression analysis are presented as hazard ratios (HRs) with corresponding 95% confidence intervals (CIs).

## Results

### Study cohort, LVEF distribution, and classification of the HFpEF cohort

We identified 8606 patients with HF, comprising 3868 HFrEF patients (LVEF < 40%), 1011 HFmrEF patients (LVEF between 40% and 49%) and 3727 with HF and LVEF ≥ 50% (**Figure 1a**). The demographics, clinical characteristics and echocardiogram data of the groups are displayed in **Supplementary Table 1**. Of the 3727 patients with HF and LVEF ≥ 50%, 311 (8.3%) had confirmed HFpEF, while 2811 (75.4%) were categorised as Undiagnosed HFpEF (ESC Criteria). 347 (9.3%) were Simple criteria, while 258 (6.9%) had an alternative diagnosis (see **Figure 1b, Graphical Abstract**). Manual validation of 100 randomly selected Confirmed HFpEF patients showed 100% of them being true positive.

**Figure 1.**
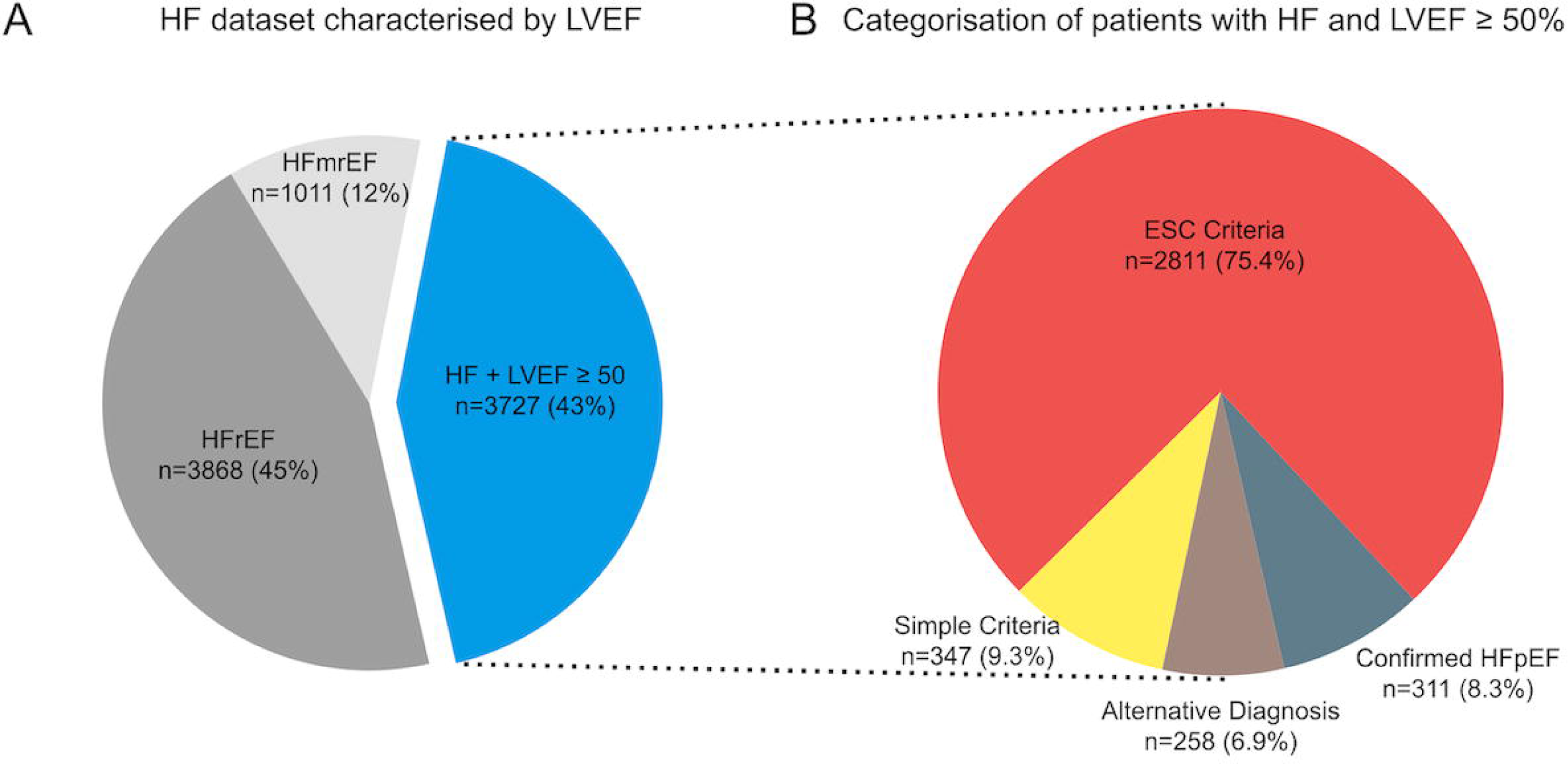
**Panel A.** The HF dataset (n=8606), categorised by LVEF. HFrEF = LVEF<40%, HFmrEF = LVEF 40-50%. **Panel B**: Patients with HF and LVEF ≥ 50% (n=3727), categorised by Confirmed HFpEF, Undiagnosed HFpEF (ESC criteria), Simple criteria, or Alternative Diagnosis.

The Confirmed HFpEF group had an average H_2_FPEF score of 6 [4, 7] (median [Interquartile range (IQR)]), which was significantly higher than both the Undiagnosed HFpEF (ESC Criteria) and the Simple Criteria groups (P<0.001 for both comparisons, **Figure 2**). The Undiagnosed HFpEF (ESC Criteria) group had a median H_2_FpEF score of 4 [3,6] (median [IQR]), which was significantly higher than the Simple Criteria group (P<0.001) which had a median H_2_FpEF score of 2 (1, 4) (median [IQR]).

**Figure 2.**
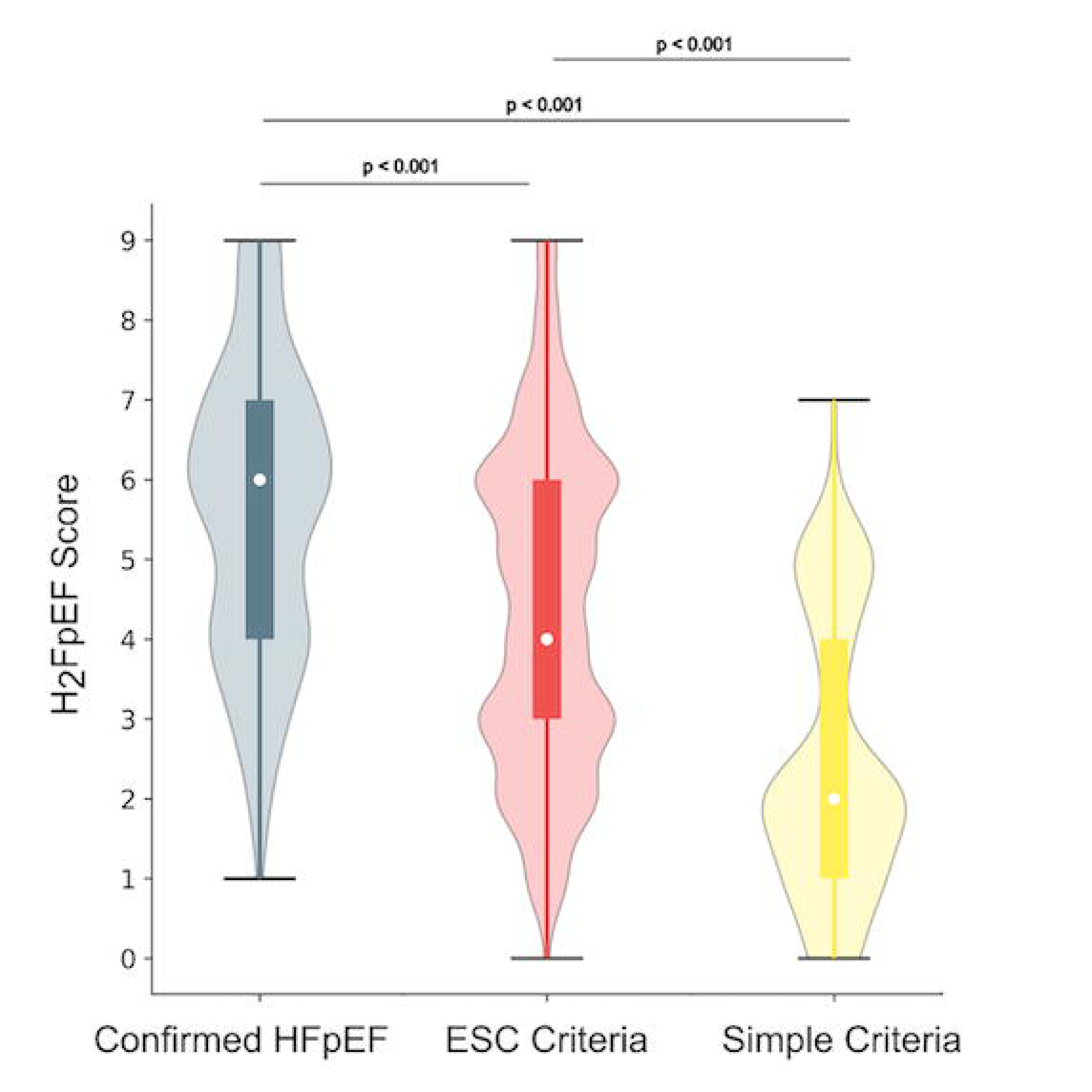
H_2_FpEF scores for subgroups, as analysed by Mann-Whitney U test corrected for multiple testing. Data are displayed as median [IQR].

**Table 1** shows a comparison of demographic and clinical characteristics between the HF and LVEF ≥ 50% subgroups. Within the Undiagnosed HFpEF (ESC Criteria) group, the most-commonly identified criterion was TR velocity >2.8mm/sec (n=2212). **Table 2** shows the number of patients meeting the individual criteria. **Table 3** shows comparison of the ESC Criteria between the subgroups.

**Table 1.** Demographic and clinical characteristic between the 3 subgroups.

|  | **Group 1** | **Group 2** | **Group 3** | **P value** |
| --- | --- | --- | --- | --- |
|  | **Confirmed**<br>**HFpEF**<br>**(n=311)** | **Undiagnosed**<br>**HFpEF (ESC**<br>**criteria)**<br>**(n=2811)** | **Simple criteria)**<br>**(n=347)** |  |
| **Demographic** |  |  |  |  |
| Female | 65% | 58% | 50% | 0.0004 |
| Age at first HF mention | 77 ± 10 | 72 ± 15 | 66 ± 17 | <0.0001 |
| Ethnicity (White) | 56% | 60% | 62% | 0.2466 |
| Ethnicity (Black) | 26% | 23% | 18% | 0.0525 |
| Ethnicity (Asian) | 9% | 5% | 5% | 0.0398 |
| **Comorbidities** |  |  |  |  |
| Diabetes type 1 | 16% | 13% | 11% | 0.2167 |
| Diabetes type 2 | 42% | 35% | 25% | <0.0001 |
| Ischaemic heart disease | 51% | 45% | 41% | 0.0388 |
| Stroke | 46% | 38% | 35% | 0.0074 |
| Hypertension | 95% | 84% | 76% | <0.0001 |
| Transient ischaemic<br>attack | 27% | 18% | 21% | 0.0003 |
| Atrial fibrillation | 57% | 43% | 29% | <0.0001 |
| Pulmonary hypertension | 35% | 28% | 12% | <0.0001 |
| Chronic kidney disease | 77% | 63% | 44% | <0.0001 |
| Myocardial infarction | 24% | 19% | 15% | 0.0085 |
| Depressive disorder | 35% | 34% | 33% | 0.7934 |
| Number of comorbidities | 4.2 ± 1.6 | 3.5 ± 1.7 | 2.7 ± 1.8 | <0.0001 |
| **Symptoms** |  |  |  |  |
| Breathlessness (NYHA II-III) | 96% | 92% | 84% | <0.0001 |
| Breathlessness (NYHA IV) | 9% | 6% | 3% | 0.0068 |
| Chest pain | 71% | 62% | 56% | 0.0004 |
| Dizziness | 58% | 41% | 36% | <0.0001 |
| Presyncope | 4% | 4% | 3% | 0.4325 |
| Syncope | 21% | 17% | 16% | 0.1564 |
| **Lifestyle** |  |  |  |  |
| Non-smoker | 86% | 72% | 58% | <0.0001 |
| **Medications** |  |  |  |  |
| Calcium channel blocker | 68% | 53% | 40% | <0.0001 |
| ACEi/ARB | 80% | 68% | 61% | <0.0001 |
| Beta-blocker | 74% | 63% | 49% | <0.0001 |
| Loop diuretic | 92% | 78% | 58% | <0.0001 |
| Statin | 70% | 52% | 43% | <0.0001 |
| Metformin | 31% | 25% | 21% | 0.0178 |
| Antiplatelet agent | 67% | 61% | 55% | 0.0075 |
| Anticoagulant | 61% | 53% | 35% | <0.0001 |
| Mineralocorticoid<br>receptor antagonist | 48% | 27% | 18% | <0.0001 |
| Angiotensin<br>receptor/neprilysin<br>inhibitor | 3% | 1% | 1% | 0.0204 |
| SGLT2 | 2% | 2% | 2% | 0.7283 |
| **Cardiology consultations** |  |  |  |  |
| Cardiac letters after first<br>HF mention | 3 (5) | 2 (3) | 1 (1) | <0.0001 |

The mean ± SD (standard deviation) are reported for variables follow a normal distribution, whereas for variables with a skewed distribution, the median (interquartile range, 25th - 75th) are reported. To evaluate the distributional differences between the groups, ANOVA test or Kruskal-Wallis H-test was used where appropriate for continuous variables while Chi-squared test was used for categorical variables. Due to the explorative and summary nature of the table, no correction for multiple comparisons has been applied.

**Table 2.** No. of undiagnosed HFpEF patients meeting the ESC Criteria.

| Criteria | No. of undiagnosed HFpEF patients |
| --- | --- |
| LV mass index (Female $\geq 95 \text{ g/m}^2$, Male $\geq 115 \text{ g/m}^2$) | 372 |
| LV mass (>219g in males and >173g in females) | 1075 |
| LA volume index (> 34 ml/m<sup>2</sup> if no AF, >40 ml/m<sup>2</sup> if AF) | 248 |
| LA volume (>58ml in males and >52ml in females) | 1114 |
| E/e' average (>9) | 895 |
| NT-proBNP (>125 for no AF, >365 for AF) | 909 |
| BNP (>35 for no AF, >105 for AF) | 764 |
| TR velocity (>2.8 m/s) | 2212 |
| **At least 1 of above** | **2811** |

**Table 3.**
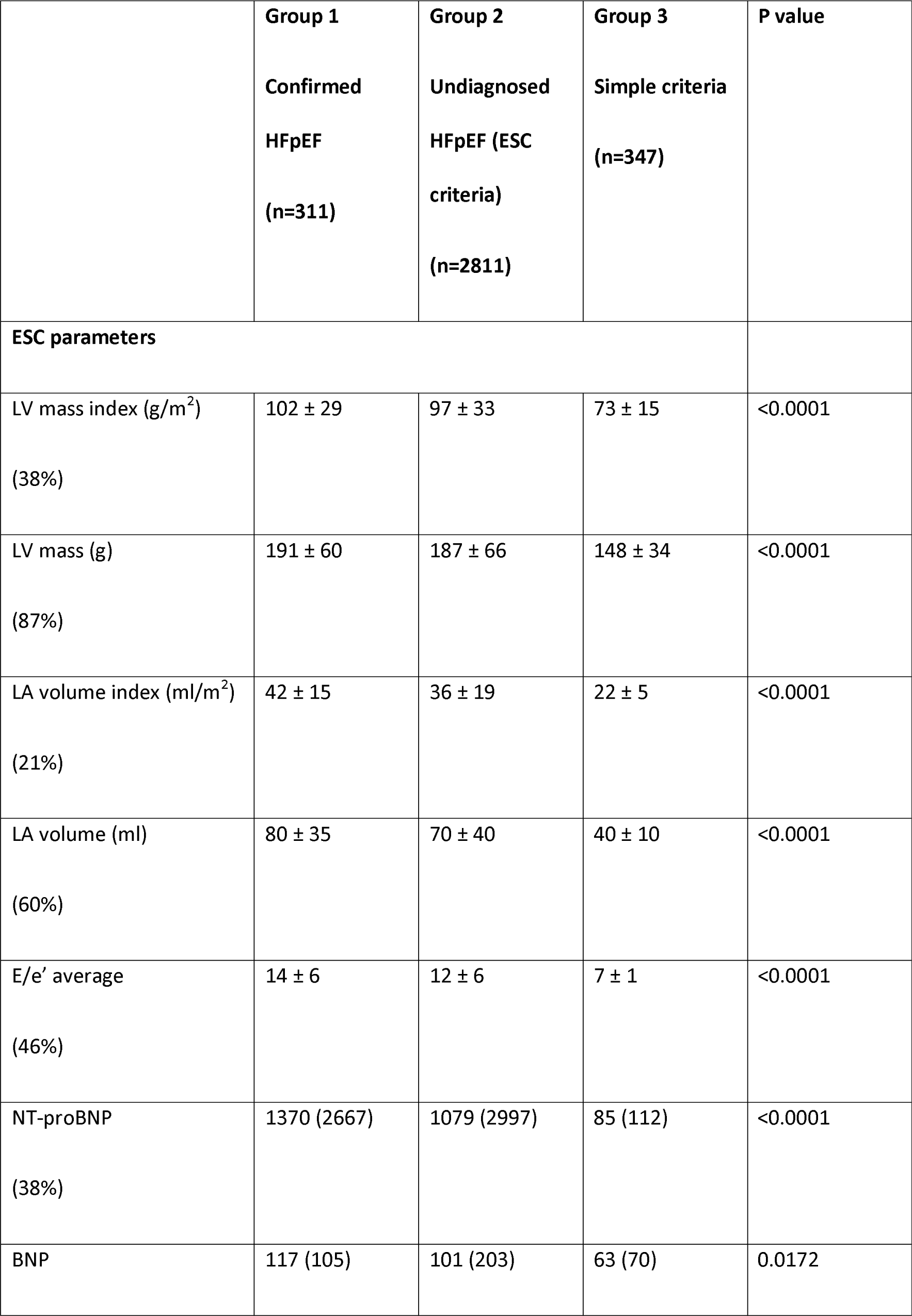
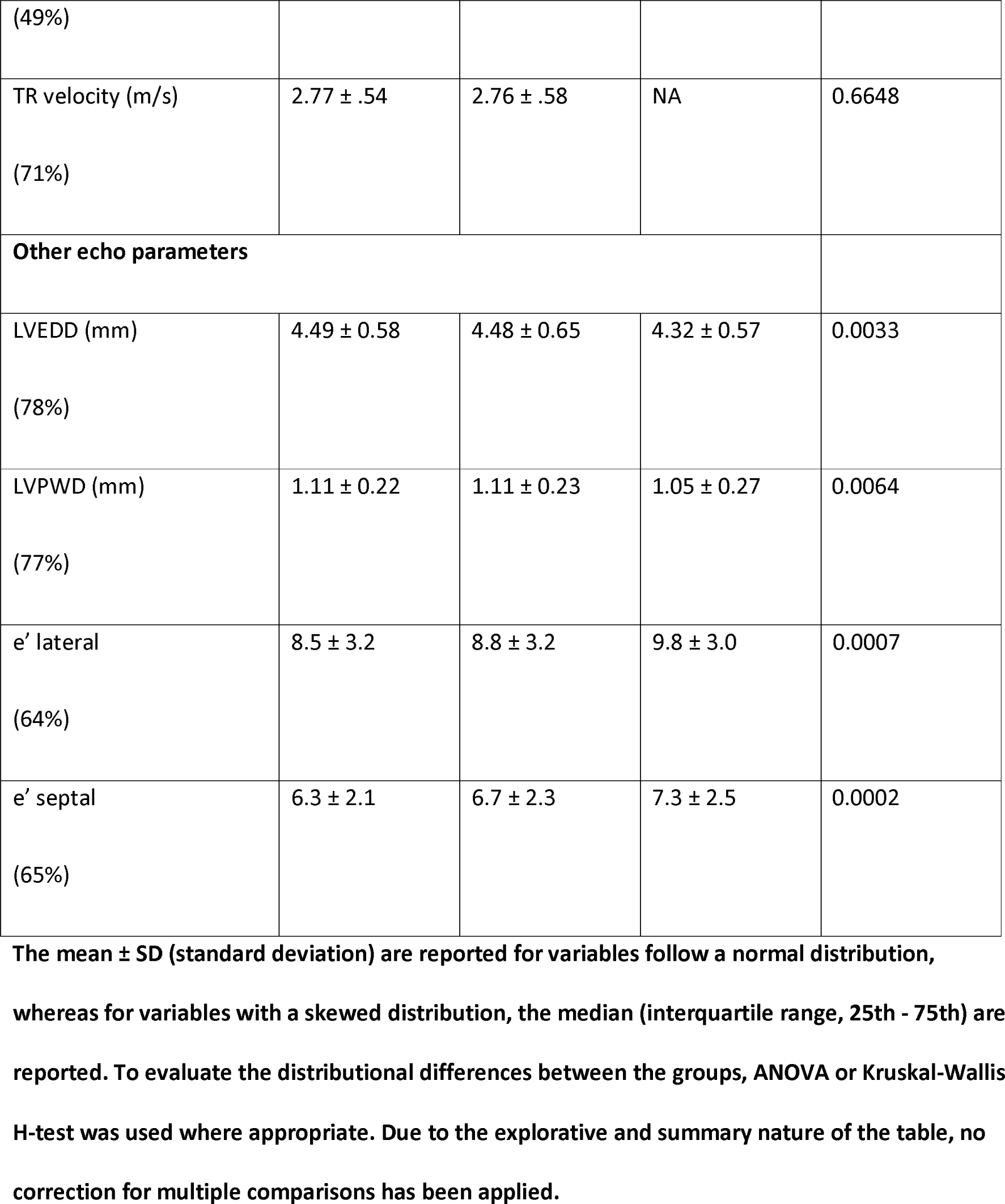
ESC parameters between the 3 subgroups.

### Clinical Outcomes

The Confirmed HFpEF group had significantly more hospitalisations within 5 years of the first HF mention in the EHR than the Undiagnosed HFpEF (ESC Criteria) (P=0.0255) and Simple criteria groups (P<0.001), **Figure 3**. We measured cardiovascular complications and found that the Confirmed HFpEF group had a significantly higher incidence of stroke and myocardial infarction than the Undiagnosed HFpEF (ESC Criteria) and Simple Criteria groups, **Figure 4**. However, when we assessed all-cause mortality, the Undiagnosed HFpEF (ESC criteria) group had significantly lower survival when compared to the Confirmed HFpEF (P=0.005) and the Simple Criteria group (P=0.002), **Figure 5a**. These findings remained significant when we compared only those patients with a H_2_FPEF score of ≥ 4 (**Supplementary Figure 1**) and when we excluded patients from the Undiagnosed HFpEF (ESC Criteria) group in whom TR velocity was the only ESC criterion met (**Supplementary Figure 2**). We also identified patients with EF≥50% and elevated natriuretic peptides (**Supplementary Figure 3**). This approach identified 942 patients, therefore accounting for approximately one third of the Undiagnosed HFpEF group. The “EF ≥50% and elevated natriuretic peptides” group had similar outcomes to the Confirmed HFpEF group in terms of hospitalisation and mortality.

**Figure 3.**
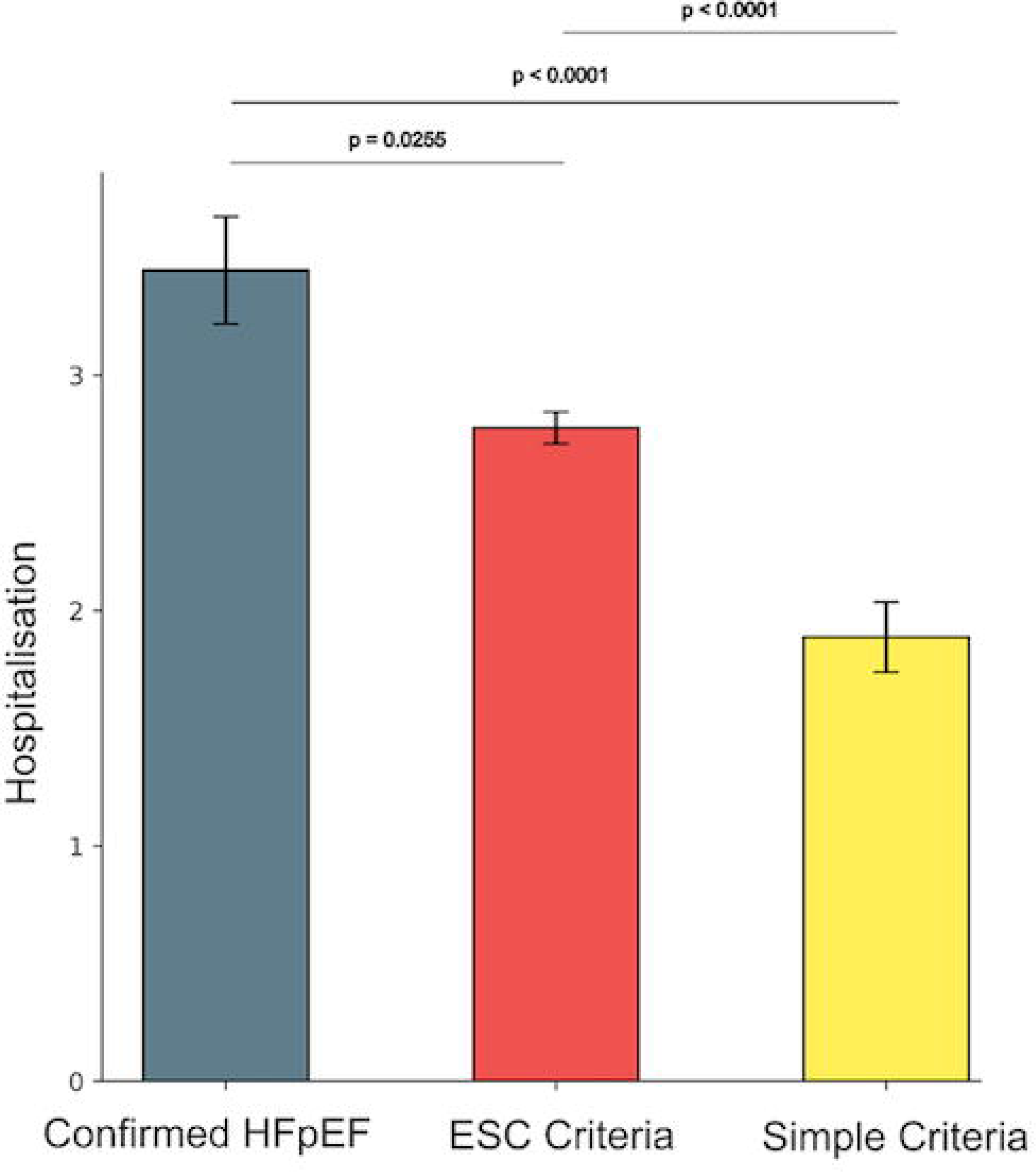
Hospitalisations after first mention of HF. Average number of hospitalisations within 5 years after first mention of HF.

**Figure 4.**
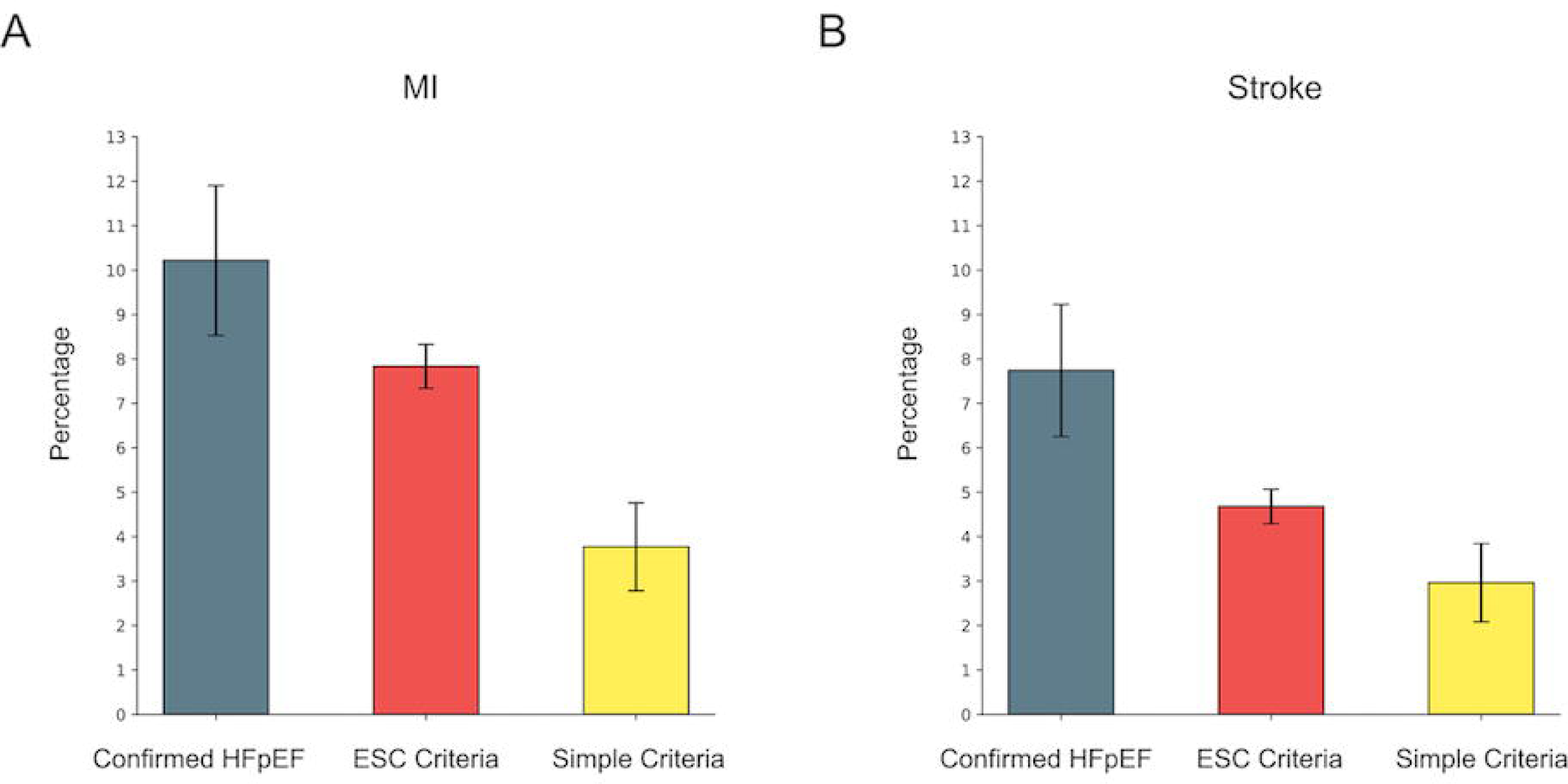
MI/Stroke percentage after first mention of HF. Data presented as mean ± SEM.

**Figure 5.**
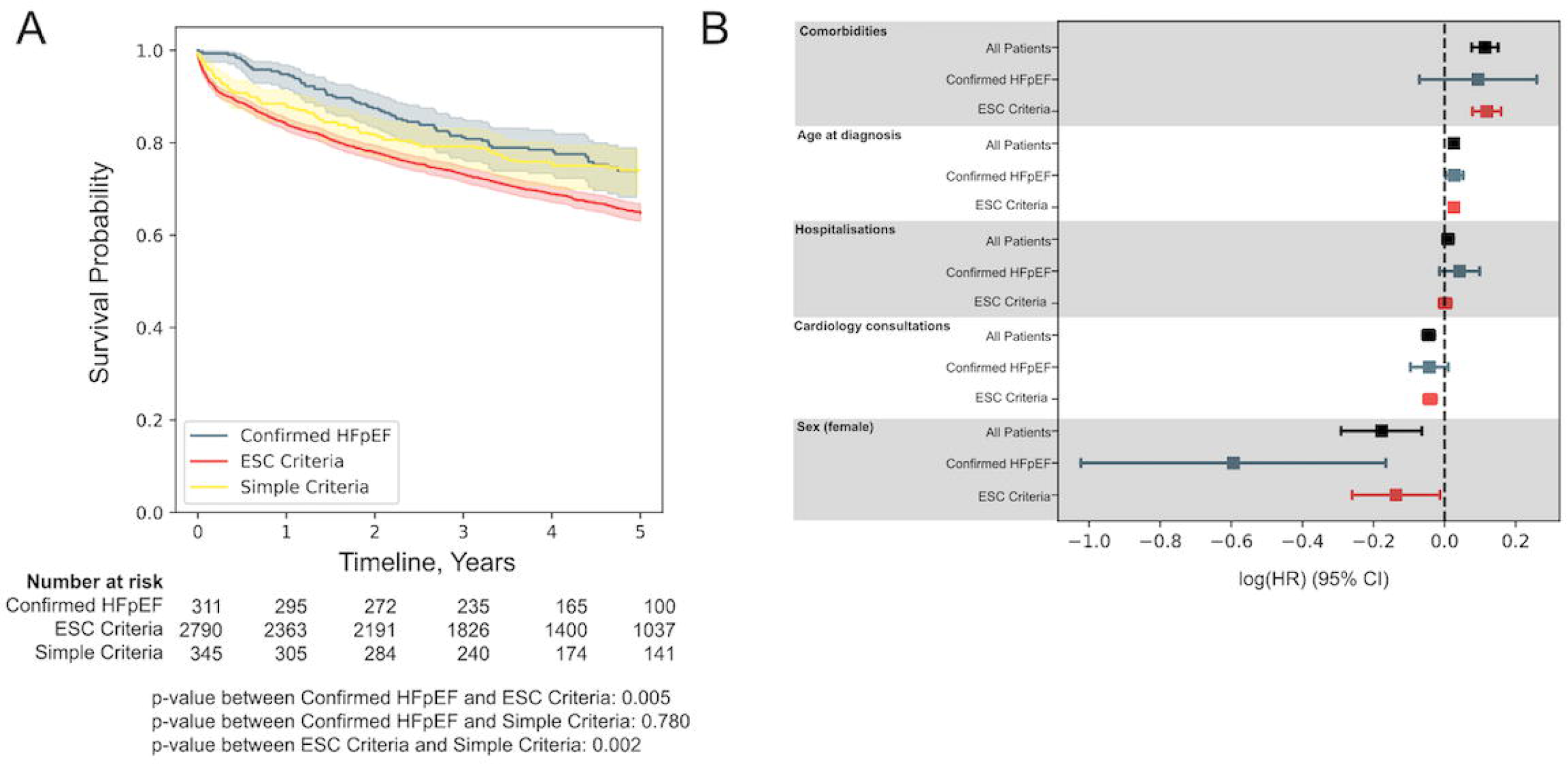
KM curves for all-cause mortality after first mention of HF, and Cox regression coefficients of various covariates.

We found that increasing age and comorbidity burden were associated with increased mortality, while female sex and seeing a cardiologist were protective features. The number of hospitalisations was not significantly associated with an increased risk of death (**Figure 5b**).

### External validation

The validation dataset (RBH cohort) consisted of 1765 with HF and LVEF >50% (53% of the total HF cohort). 173 (10%) patients had Confirmed HFpEF, while 131 (7%) had an alternative diagnosis and 1461 (83%) had undiagnosed HFpEF (it was not possible to fully characterise the undiagnosed HFpEF group due to lack of data on ESC criteria, **Supplementary Table 3, Supplementary Figure 4**. We identified a similar pattern of clinical characteristics as in the KCHNFT cohort. Moreover, a similar difference in mortality was seen between Confirmed HFpEF and undiagnosed HFpEF. In the RBH cohort, age, comorbidities and hosptialisations were associated with increased mortality, while female sex and seeing a cardiologist were protective features.

### Temporal analysis

Over the study period, the proportion of patients diagnosed with confirmed HFpEF versus Undiagnosed HFpEF (ESC Criteria) over a 3 year rolling average increased from 2% (2010-2012 inclusive) to 16% (2018-2020 inclusive). Supplementary Figure 5 shows the trend of the confirmed HFpEF and Undiagnosed HFpEF (ESC Criteria) by years.

## Discussion

In this study we characterised a real-world HF population and looked in detail at those patients with features of HFpEF. We found that 91% of patients with HF and an LVEF>50% on echocardiogram did not have a formal diagnosis of HFpEF. Of the patients with undiagnosed HFpEF, 75% had clinical characteristics to satisfy a formal diagnosis of HFpEF as per the ESC diagnostic criteria. Despite being younger and less comorbid, the Undiagnosed HFpEF (ESC Criteria) group had similar rates of hospitalisation to those with Confirmed HFpEF and had a significantly worse 5-year survival.

It is likely that the increased mortality in the Undiagnosed HFpEF (ESC Criteria) group is related to a combination of factors. Despite being younger and less comorbid, this group were less likely to be seen by a cardiologist and there was lower utilisation of HF medications, both of which are important in the prognosis of HF.^17,18^ In addition, the Undiagnosed HFpEF (ESC Criteria) group had a higher proportion of male patients than the Confirmed HFpEF group, while female sex was noted to be a factor associated with improved survival: it is possible that this represents underlying sex bias in the diagnosis of HFpEF. Regardless, the finding that the Undiagnosed HFpEF (ESC Criteria) group have a higher mortality highlights the need for accurate diagnosis so this group can derive benefit from evidence-based HFpEF management and therapies.

It is unclear why patients with clinical features meeting the ESC Criteria for HFpEF remain without a formal diagnosis. While patients with Undiagnosed HFpEF (ESC Criteria) had a lower median H2FPEF score than those with Confirmed HFpEF (4 vs 6), even a score of 4 suggests a likelihood of HFpEF of >70%. It is likely that the lower frequency of cardiology consultation in the Undiagnosed HFpEF (ESC Criteria) group is important given previous evidence that non-cardiologists and primary care physicians are less likely to diagnose HFpEF than cardiology specialists. ^19^ This highlights a weakness of points-based diagnostic algorithms such as H2FPEF score; they rely on an *a priori* suspicion of HFpEF diagnosis upon clinical assessment or may not even be used widely by treating physicians. User friendly machine learning techniques therefore have potential to complement existing diagnostic algorithms via automated identification of suspected/likely HFpEF who can then undergo formal diagnostic assessment by an expert clinician. Although we have used an approach that focussed on structured and unstructured data within the EHR, other AI-based approaches using cardiac imaging data (e.g. echocardiographic images) also demonstrate an ability to discriminate between HFpEF and non-HFpEF and are of high potential for development. ^20^It is notable that the undiagnosed HFpEF (ESC Criteria) and Simple criteria groups have significantly different clinical characteristic, echo findings, clinical complications and all-cause mortality. The median H_2_FPEF score of the Simple Criteria group is low: 2, suggesting a likelihood of HFpEF of <40%, while this group have no echocardiographic or biochemical features to suggest diastolic dysfunction. However, even if it assumed that this group do not in fact have HFpEF at all, at least not in the contemporary understanding of HFpEF, one must question why these patients have presented with symptoms and/or signs of HF. It is possible that this group have been misdiagnosed as HF e.g. with the true diagnosis being another cause of breathlessness, such as pulmonary disease with or without *cor pulmonale*.

The findings of this study may have implications for clinical trial design. Many previous randomised control trials of potential HFpEF therapies have used a LVEF >40% as the sole criterion to be included in the HFpEF group. Our data suggests that such an approach will result in approximately 20% of the ‘HFpEF’ patients enrolled in such a trial not having clinical features to support a diagnosis of HFpEF. Such a recruitment strategy would therefore confer an increased risk of Type 1 error. Moreover, HFpEF is widely considered to be a heterogenous population, but considering the broad similarities between the Confirmed HFpEF and Undiagnosed HFpEF (ESC Criteria) groups in this study, perhaps at least some of this heterogeneity comes from overly lax inclusion criteria in clinical trial design rather than true clinical heterogeneity within the HFpEF population.

The strengths of our study include the use of NLP to identify HF mentions in the clinical text rather than relying on formal clinical coding. Our approach has been validated in a number of other disease settings in other studies and is supported by manual validation of randomly selected HFpEF patients from within this study. A further strength of our approach is that it includes data from both an inpatient and outpatient hospital setting and does not rely on HF hospitalisation to identify patients. Our Cogstack informatics platform also allows us to incorporate structured and unstructured data from the EHR, including echocardiographic data, clinical complications and mortality data. Finally, our results were validated in a second independent cohort.

### Limitations

This study is retrospective and is therefore prone to the same biases as other observational studies. Our study findings should be considered only as hypothesis generating only. Hospitalisations only apply to our centre and may miss admissions to other hospitals. Although we had good data on mortality, cause of death was not available, nor was cause of hospitalisation (e.g. heart failure hospitalisation versus other causes). During characterisation of the Undiagnosed HFpEF (ESC Criteria) group, it was apparent that there were very few recorded values for pulmonary artery systolic pressure in the echocardiogram reports. We therefore had to rely solely on a tricuspid regurgitation velocity of >2.8 m/s instead of PASP for this criterion. No exericse data was collected in this study.

### Conclusion

In this study we identify that there may be a substantially larger than thought caseload of clinical HFpEF that remains without a formal diagnosis. NLP algorithm-based identification of these patients represents a promising approach. Patients with HFpEF who lack a formal diagnosis are younger and less co-morbid but have a worse prognosis than those with a clinician-assigned diagnosis of HFpEF.

### Funding

This work was supported by grants from the British Heart Foundation (CH/1999001/11735, RG/20/3/34823 and RE/18/2/34213 to AMS; CC/22/250022 to RJDB, AMS, JT and KOG) and King’s College Hospital Charity (D3003/122022/Shah/1188 to AMS). KOG and DIB are each supported by MRC Clinician Scientist Fellowships (MR/Y001311/1 to KOG, MR/X001881/1 to DiB).

## Data Availability

The datasets analysed during the current study are not publicly available due to hospital information governance regulations but are available from the corresponding author on reasonable request.

## Supplementary Figures

**Supplementary Figure 1.**
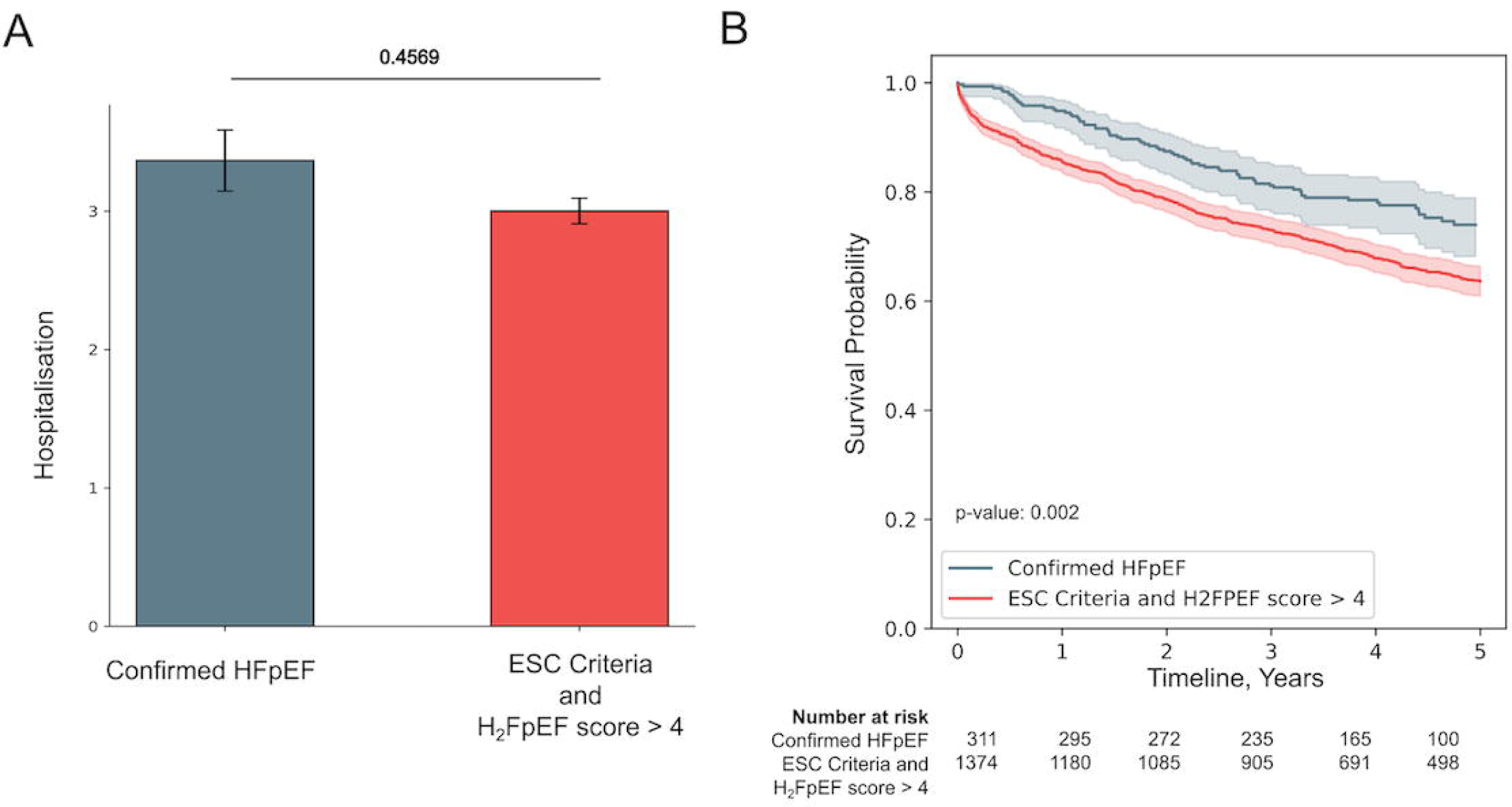
Comparison between Confirmed HFpEF group and Undiagnosed HFpEF (ESC criteria) with H2FPEF score > 4 (n=1379). Panel A: Hospitalisations within 5 years. Panel B: KM curve for all-cause mortality.

**Supplementary Figure 2.**
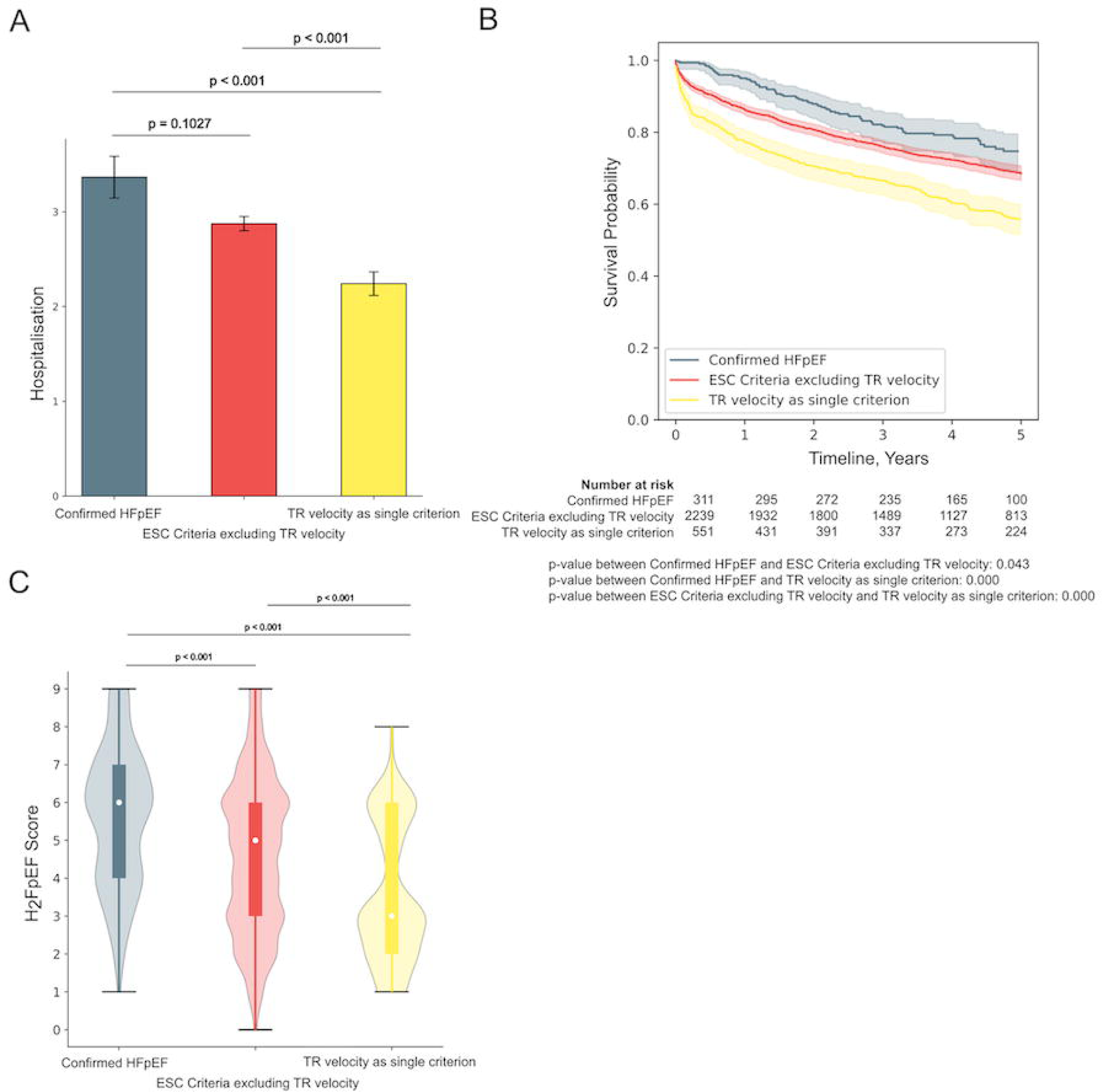
Comparison between Confirmed HFpEF group and Undiagnosed HFpEF (ESC criteria) excluding patients with satisfying only the TR velocity criteria (561 patients excluded, leaving n=2250). Panel A: Hospitalisations within 5 years. Panel B: KM curve for all-cause mortality. Panel C: Comparison of H2FPEF score of the 3 groups.

**Supplementary Figure 3.**
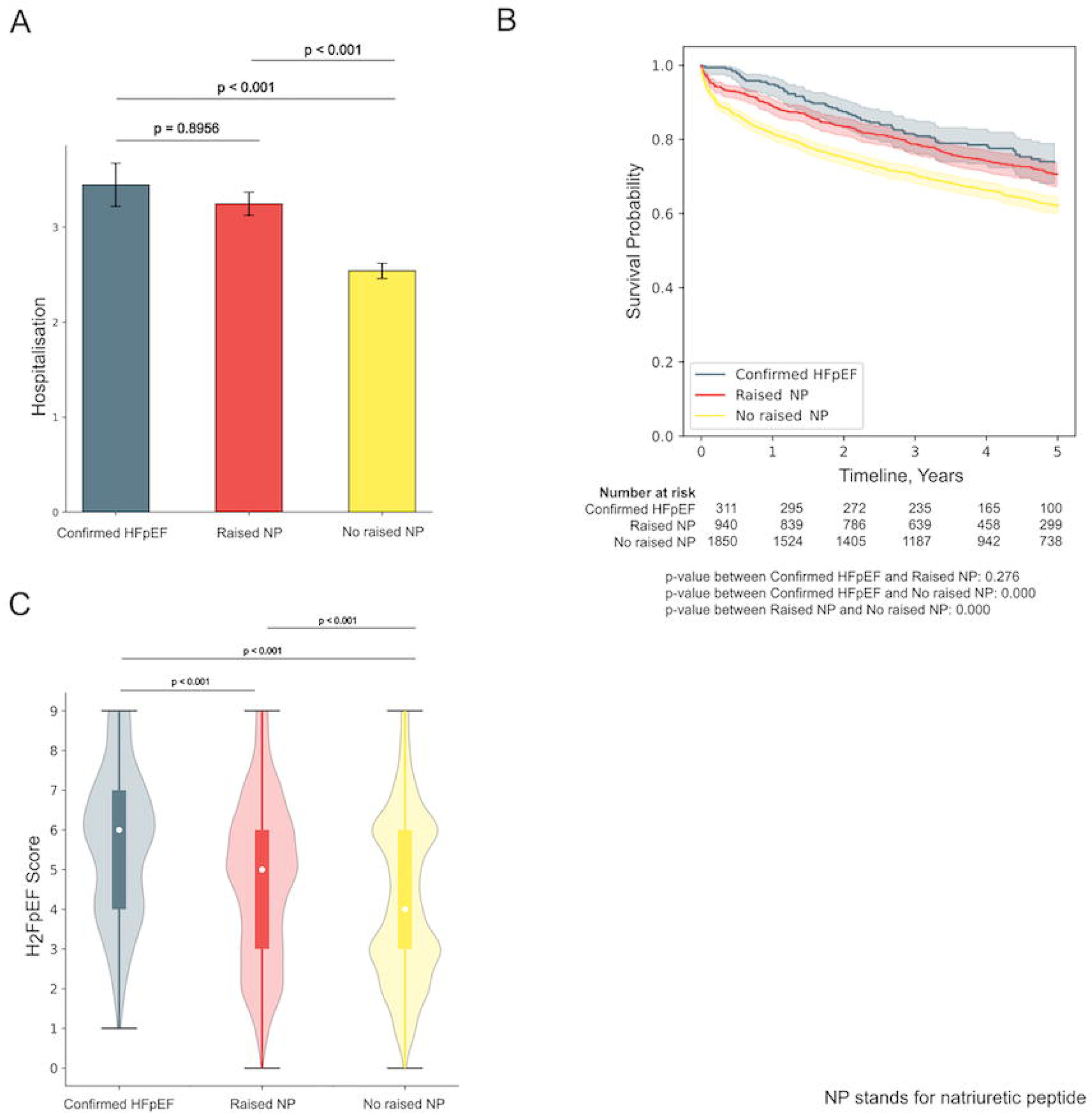
Comparison between Confirmed HFpEF group and patients with raised NP (natriuretic peptide) in the Undiagnosed HFpEF (ESC criteria) group (n=942), the remaining patients without raised NP (n=1869) are placed into the ‘No raised NP’ group. Panel A: Hospitalisations within 5 years. Panel B: KM curve for all-cause mortality. Panel C: Comparison of H2FPEF score of the 3 groups.

**Supplementary Figure 4.**
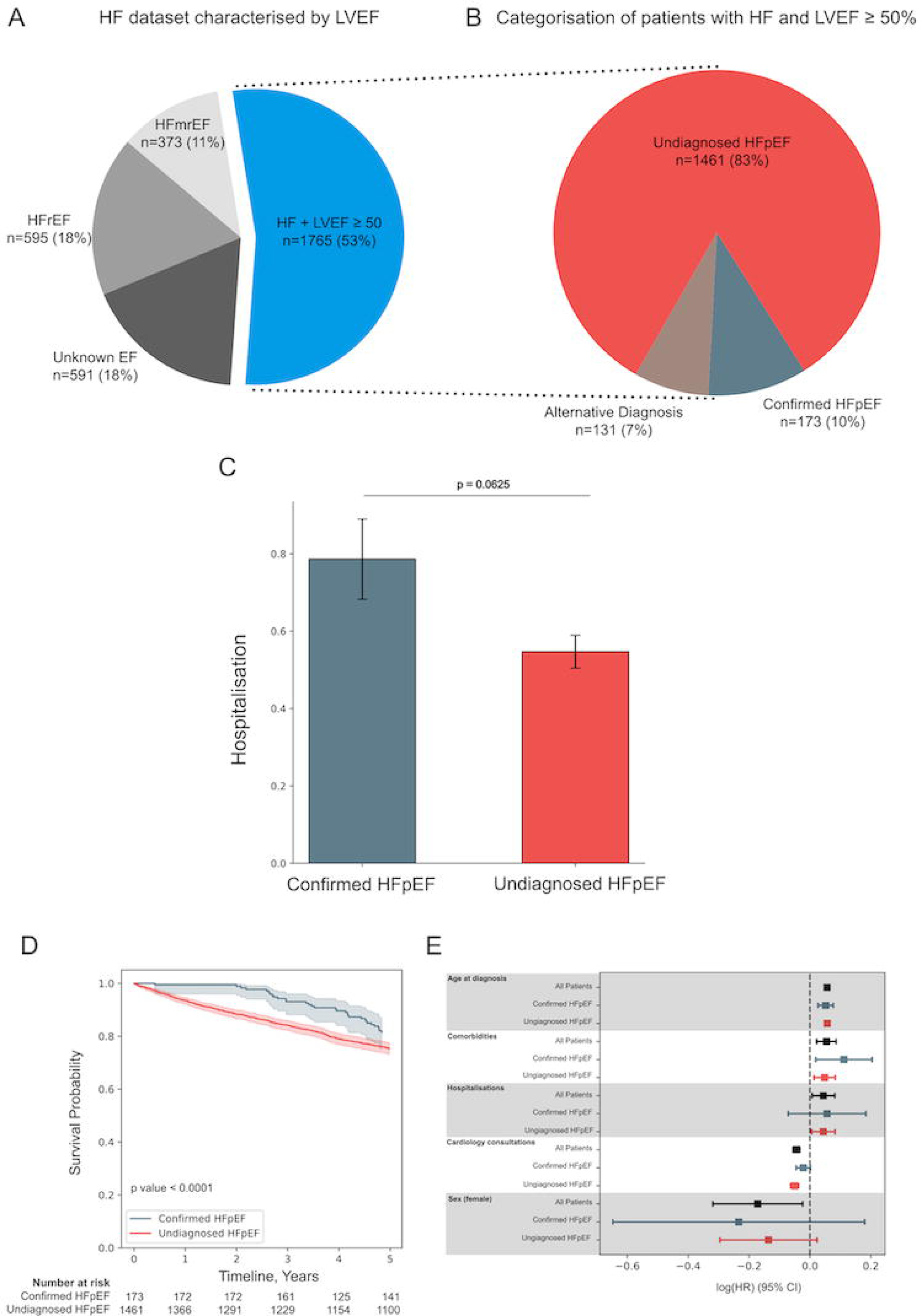
Results of external validation using the RBH cohort. Panel A. The HF dataset (n=3324), categorised by LVEF. HFrEF = LVEF<40%, HFmrEF = LVEF 40-50%. Panel B: Patients with HF and LVEF ≥ 50% (n=1765), categorised by Confirmed HFpEF, Undiagnosed HFpEF or Alternative Diagnosis. Panel C: Average number of hospitalisations within 5 years after first mention of HF. Panel D: KM curves for all-cause mortality after first mention of HF, and Panel E: Cox regression coefficients of various covariates.

**Supplementary Figure 5.**
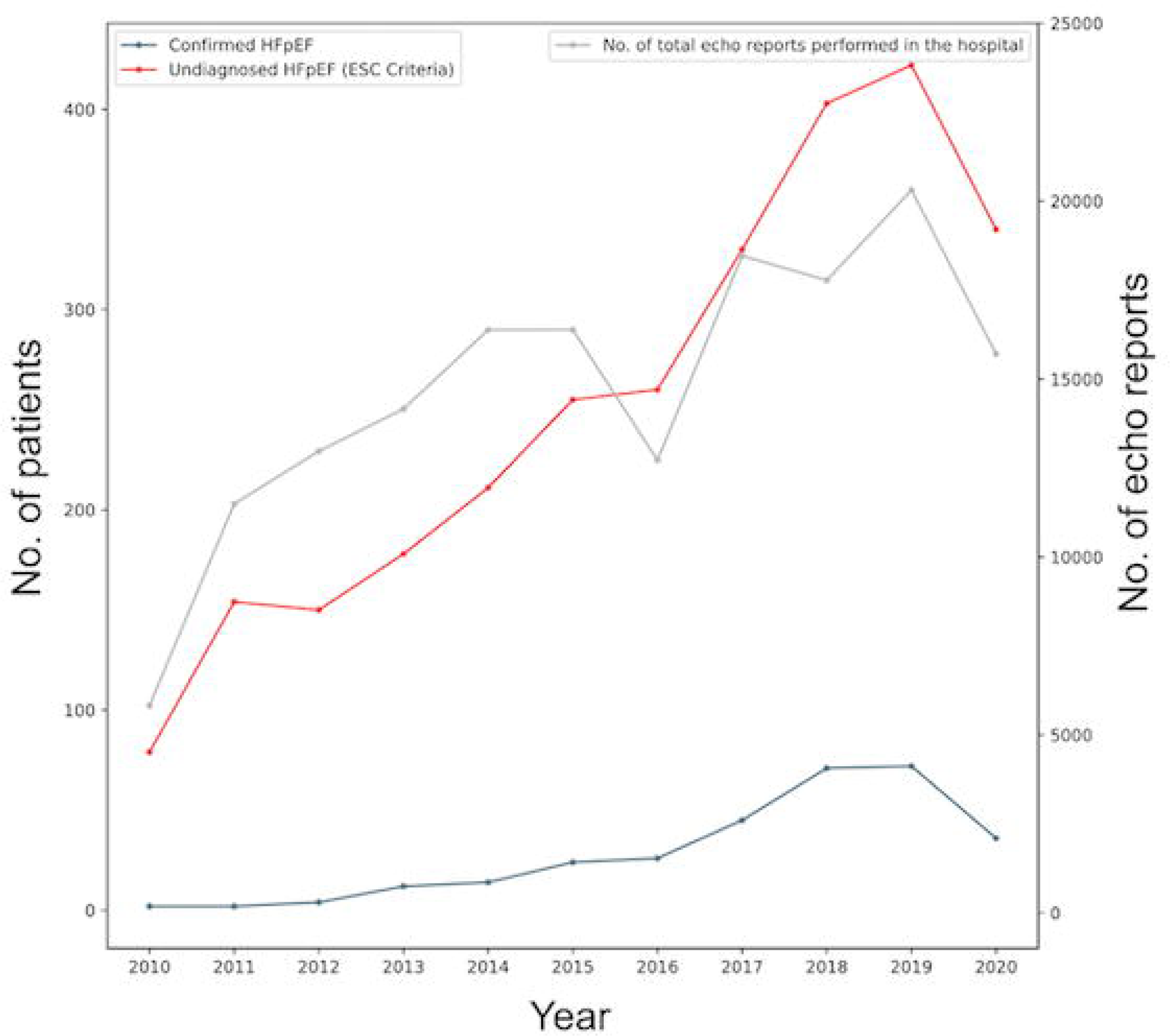
The temporal trends of the Confirmed HFpEF and Undiagnosed HFpEF by years, together with the total number of echo reports performed in the hospital.

**Supplementary Table 1.** Characteristics of HF patients categorised by LVEF.

|  | **HF and LVEF ≥**<br>**50%**<br>**(n=3727)** | **HFmrEF**<br>**(n=1011)** | **HFrEF**<br>**(n=3868)** | **P value** |
| --- | --- | --- | --- | --- |
| **Demographic data** |  |  |  |  |
| Female | 57% | 36% | 28% | <0.0001 |
| Age at first HF<br>mention | 72 ± 15 | 72 ± 14 | 68 ± 14 | <0.0001 |
| Ethnicity (White) | 59% | 68% | 63% | <0.0001 |
| Ethnicity (Black) | 23% | 14% | 20% | <0.0001 |
| Ethnicity (Asian) | 6% | 4% | 6% | 0.0239 |
| **Comorbidities** |  |  |  |  |
| Diabetes type 1 | 13% | 9% | 10% | <0.0001 |
| Diabetes type 2 | 34% | 29% | 33% | 0.0119 |
| Ischaemic heart<br>disease | 46% | 61% | 74% | <0.0001 |
| Myocardial<br>infarction | 19% | 37% | 54% | <0.0001 |
| Hypertension | 85% | 80% | 77% | <0.0001 |
| Stroke | 38% | 32% | 38% | 0.0002 |
| Transient ischaemic attack | 19% | 12% | 12% | <0.0001 |
| Atrial fibrillation | 43% | 44% | 42% | 0.2071 |
| Pulmonary hypertension | 28% | 14% | 13% | <0.0001 |
| Chronic kidney disease | 63% | 55% | 56% | <0.0001 |
| Depressive disorder | 34% | 27% | 28% | <0.0001 |
| **Lifestyle** |  |  |  |  |
| Non-smokers | 72% | 60% | 66% | <0.0001 |
| **Symptoms** |  |  |  |  |
| Breathlessness (NYHA I-III) | 92% | 89% | 92% | 0.0027 |
| Breathlessness (NYHA IV) | 6% | 4% | 4% | 0.0002 |
| Chest pain | 63% | 60% | 70% | <0.0001 |
| Dizziness | 43% | 32% | 36% | <0.0001 |
| Presyncope | 4% | 4% | 6% | 0.0001 |
| Syncope | 18% | 15% | 16% | 0.0307 |
| **Echo data** |  |  |  |  |
| LVEDD (mm) | 4.5 ± 0.6 | 4.9 ± 0.8 | 5.7 ± 0.9 | <0.0001 |
| LVPWD (mm) | 1.1 ± 0.23 | 1.1 ± 0.24 | 1.0 ± 0.25 | <0.0001 |
| LV mass (g) | 187 ± 67 | 217 ± 82 | 247 ± 83 | <0.0001 |
| LV mass index<br>(g/m<sup>2</sup>) | 99 ± 34 | 112 ± 37 | 127 ± 40 | <0.0001 |
| LA Volume (ml) | 72 ± 41 | 74 ± 41 | 80 ± 35 | <0.0001 |
| LA Volume index<br>(ml/m<sup>2</sup>) | 38 ± 19 | 37 ± 15 | 43 ± 18 | <0.0001 |
| E/E' Average | 12 ± 6 | 13 ± 6 | 14 ± 7 | <0.0001 |
| TR max vel. (m/s) | 2.76 ± 0.58 | 2.70 ± 0.54 | 2.69 ± 0.54 | <0.0001 |
| **Lab tests** |  |  |  |  |
| BMI (kg/m<sup>2</sup>) | 29 ± 8 | 28 ± 7 | 27 ± 6 | <0.0001 |
| Haemoglobin (g/dl) | 121 ± 21 | 124 ± 22 | 124 ± 22 | <0.0001 |
| BNP (pg/mL) | 105 (191) | 191 (537) | 325 (611) | <0.0001 |
| NT-pro-BNP (pg/mL) | 1171 (3005) | 1548 (3405) | 2624 (7660) | <0.0001 |
| HDL (mmol/L) | 1.3 (0.5) | 1.2 (0.4) | 1.2 (0.5) | <0.0001 |
| LDL (mmol/L) | 2.2 (1.2) | 2.1 (1.3) | 2.2 (1.3) | 0.0526 |
| Creatinine (umol/L) | 89 (51) | 93 (50) | 97 (53) | <0.0001 |
| **Medications** |  |  |  |  |
| Calcium channel | 53% | 40% | 35% | <0.0001 |
| blocker |  |  |  |  |
| ACEi/ARB | 69% | 85% | 95% | <0.0001 |
| Beta-blocker | 64% | 85% | 95% | <0.0001 |
| Loop diuretic | 78% | 74% | 83% | <0.0001 |
| Statin | 53% | 53% | 63% | <0.0001 |
| Metformin | 25% | 20% | 24% | 0.0125 |
| Antiplatelet agent | 62% | 69% | 77% | <0.0001 |
| Anticoagulant | 52% | 50% | 54% | 0.0651 |
| Mineralocorticoid<br>receptor antagonist | 29% | 46% | 74% | <0.0001 |
| Angiotensin<br>receptor/neprilysin<br>inhibitor | 1% | 2% | 5% | <0.0001 |
| SGLT2 | 2% | 4% | 10% | <0.0001 |

The mean ± SD (standard deviation) are reported for variables follow a normal distribution, whereas for variables with a skewed distribution, the median (interquartile range, 25th - 75th) are reported. To evaluate the distributional differences between the groups, ANOVA test or Kruskal-Wallis H-test was used where appropriate for continuous variables while Chi-squared test was used for categorical variables. Due to the explorative and summary nature of the table, no correction for multiple comparisons has been applied.

**Supplementary Table 2.** SNOMED-CT terms used in the study for identifying the HF + LVEF≥50% patients and the subgroups. All child terms are included by default.

| Patient Group | SNOMED-CT Term | ID |
| --- | --- | --- |
| HF patients | Heart failure (disorder) | 84114007 |
| Confirmed HFpEF patients | Heart failure with normal ejection fraction (disorder) | 446221000 |
| Alternative Diagnosis patients | Aortic valve stenosis (disorder)* | 60573004 |
|  | Aortic valve regurgitation (disorder)* | 60234000 |
|  | Mitral valve stenosis (disorder)* | 79619009 |
|  | Mitral valve regurgitation (disorder)* | 48724000 |
|  | Pulmonic valve stenosis (disorder)* | 56786000 |
|  | Pulmonic valve regurgitation (disorder)* | 91434003 |
|  | Hypertrophic cardiomyopathy (disorder) | 233873004 |
|  | Senile cardiac amyloidosis (disorder) | 16573007 |
|  | Restrictive cardiomyopathy (disorder) | 415295002 |
|  | Constrictive pericarditis (disorder) | 415295002 |

\* - the **severe** valvular heart disease patients are identified with the 'severe' keyword before the mentions of the SNOMED-CT terms.

**Supplementary Table 3.** Demographic and clinical characteristic between the 2 subgroups in the RBH cohort.

|  | **Group 1**<br>**Confirmed HFpEF**<br>**(n=173)** | **Group 2**<br>**Undiagnosed HFpEF**<br>**(n=1461)** | **P value** |
| --- | --- | --- | --- |
| **Demographic** |  |  |  |
| Female | 62% | 51% | 0.0061 |
| Age at first HF mention | 76 ± 10 | 63 ± 18 | <0.0001 |
| Ethnicity (White) | 66% | 69% | 0.4683 |
| Ethnicity (Black) | 22% | 18% | 0.1984 |
| Ethnicity (Asian) | 7% | 6% | 0.5660 |
| **Comorbidities** |  |  |  |
| Diabetes type 1 | 25% | 16% | 0.0031 |
| Diabetes type 2 | 59% | 43% | <0.0001 |
| Ischaemic heart disease | 65% | 54% | 0.0046 |
| Stroke | 20% | 12% | 0.0098 |
| Hypertension | 96% | 78% | <0.0001 |
| Transient ischaemic attack | 28% | 14% | 0.0046 |
| Atrial fibrillation | 80% | 58% | <0.0001 |
| Pulmonary hypertension | 29% | 20% | 0.0055 |
| Chronic kidney disease | 66% | 36% | <0.0001 |
| Myocardial infarction | 43% | 34% | 0.0188 |
| Depressive disorder | 40% | 24% | <0.0001 |
| Number of comorbidities | 6.4 ± 2.1 | 4.6 ± 2.4 | <0.0001 |
| **Symptoms** |  |  |  |
| Breathlessness (NYHA I-III) | 99% | 87% | <0.0001 |
| Breathlessness (NYHA IV) | 27% | 7% | <0.0001 |
| Chest pain | 74% | 64% | 0.0141 |
| Dizziness | 67% | 52% | 0.0002 |
| Presyncope | 13% | 9% | 0.0953 |
| Syncope | 33% | 27% | 0.1112 |
| **Lifestyle** |  |  |  |
| Non-smoker | 93% | 79% | <0.0001 |
| **Medications** |  |  |  |
| Calcium channel blocker | 68% | 47% | <0.0001 |
| ACEi/ARB | 88% | 70% | <0.0001 |
| Beta-blocker | 84% | 67% | <0.0001 |
| Loop diuretic | 95% | 54% | <0.0001 |
| Statin | 83% | 63% | <0.0001 |
| Metformin | 31% | 20% | 0.0025 |
| Antiplatelet agent | 80% | 62% | <0.0001 |
| Anticoagulant | 74% | 48% | <0.0001 |
| Mineralocorticoid<br>receptor antagonist | 49% | 26% | <0.0001 |
| Angiotensin<br>receptor/neprilysin<br>inhibitor | 2% | 2% | 1.0000 |
| SGLT2 | 4% | 4% | 0.9497 |
| **Cardiology consultations** |  |  |  |
| Cardiac letters after first<br>HF mention | 9 (12) | 5 (10) | <0.0001 |

The mean ± SD (standard deviation) are reported for variables follow a normal distribution, whereas for variables with a skewed distribution, the median (interquartile range, 25th - 75th) are reported. To evaluate the distributional differences between the groups, ANOVA test was used for continuous variables while Chi-squared test was used for categorial variables. Due to the explorative and summary nature of the table, no correction for multiple comparisons has been applied.

## References

1. Owan TE, Hodge DO, Herges RM, Jacobsen SJ, Roger VL, Redfield MM. Trends in prevalence and outcome of heart failure with preserved ejection fraction. N Engl J Med. 2006;355(3):251–259.

2. Kitzman DW, Gardin JM, Gottdiener JS, et al. Importance of heart failure with preserved systolic function in patients≥ 65 years of age. Am J Cardiol. 2001;87(4):413–419.

3. Borlaug BA, Paulus WJ. Heart failure with preserved ejection fraction: Pathophysiology, diagnosis, and treatment. Eur Heart J. 2011;32(6):670–679.

4. McDonagh TA, Metra M, Adamo M, et al. 2021 ESC guidelines for the diagnosis and treatment of acute and chronic heart failure: Developed by the task force for the diagnosis and treatment of acute and chronic heart failure of the european society of cardiology (ESC) with the special contribution of the heart failure association (HFA) of the ESC. Eur Heart J. 2021;42(36):3599–3726.

5. Reddy YN, Carter RE, Obokata M, Redfield MM, Borlaug BA. A simple, evidence-based approach to help guide diagnosis of heart failure with preserved ejection fraction. Circulation. 2018;138(9):861–870.

6. Pieske B, Tschöpe C, De Boer RA, et al. How to diagnose heart failure with preserved ejection fraction: The HFA–PEFF diagnostic algorithm: A consensus recommendation from the heart failure association (HFA) of the european society of cardiology (ESC). Eur Heart J. 2019;40(40):3297–3317.

7. Anker SD, Butler J, Filippatos G, et al. Empagliflozin in heart failure with a preserved ejection fraction. N Engl J Med. 2021;385(16):1451–1461.

8. Nassif ME, Windsor SL, Borlaug BA, et al. The SGLT2 inhibitor dapagliflozin in heart failure with preserved ejection fraction: A multicenter randomized trial. Nat Med. 2021;27(11):1954–1960.

9. Bean DM, Kraljevic Z, Searle T, et al. AngiotensinDconverting enzyme inhibitors and angiotensin II receptor blockers are not associated with severe COVIDD19 infection in a multiDsite UK acute hospital trust. European journal of heart failure. 2020;22(6):967–974.

10. Kraljevic Z, Searle T, Shek A, et al. Multi-domain clinical natural language processing with medcat: The medical concept annotation toolkit. Artif Intell Med. 2021;117:102083.

11. Wu H, Wang M, Wu J, et al. A survey on clinical natural language processing in the united kingdom from 2007 to 2022. NPJ digital medicine. 2022;5(1):186.

12. Farajidavar N, O’Gallagher K, Bean D, et al. Diagnostic signature for heart failure with preserved ejection fraction (HFpEF): A machine learning approach using multi-modality electronic health record data. BMC Cardiovascular Disorders. 2022;22(1):1–13.

13. Jackson R, Kartoglu I, Stringer C, et al. CogStack-experiences of deploying integrated information retrieval and extraction services in a large national health service foundation trust hospital. BMC medical informatics and decision making. 2018;18(1):1–13.

14. Searle T, Kraljevic Z, Bendayan R, Bean D, Dobson R. MedCATTrainer: A biomedical free text annotation interface with active learning and research use case specific customisation. arXiv preprint arXiv:1907.07322. 2019.

15. Johnson AE, Pollard TJ, Shen L, et al. MIMIC-III, a freely accessible critical care database. Scientific data. 2016;3(1):1–9.

16. Bean DM, Kraljevic Z, Shek A, Teo J, Dobson RJ. Hospital-wide natural language processing summarising the health data of 1 million patients. PLOS Digital Health. 2023;2(5):e0000218.

17. Lindberg F, Lund LH, Benson L, et al. Patient profile and outcomes associated with followDup in specialty vs. primary care in heart failure. ESC Heart Failure. 2022;9(2):822–833.

18. Jensen J, Poulsen MK, Petersen PW, Gerdes B, Rossing K, Schou M. Prevalence of heart failure phenotypes and current use of therapies in primary care: Results from a nationwide study. ESC Heart Failure. 2023.

19. Hancock HC, Close H, Fuat A, Murphy JJ, Hungin APS, Mason JM. Barriers to accurate diagnosis and effective management of heart failure have not changed in the past 10 years: A qualitative study and national survey. BMJ open. 2014;4(3):e003866.

20. Akerman AP, Porumb M, Scott CG, et al. Automated echocardiographic detection of heart failure with preserved ejection fraction using artificial intelligence. JACC: Advances. 2023;2(6):100452.

